# Left atrial strain tracks abnormal ventricular mechanics in Fabry disease

**DOI:** 10.1101/2025.03.02.25323185

**Authors:** Chosita Cheepvasarach, Michael Gribble, Ravi Vijapurapu, Sabrina Nordin, Joao Augusto, Martin Ugander, Richard Steeds, Michel Tchan, James C Moon, Faraz Pathan, Rebecca Kozor

## Abstract

**Backgrounds:** Fabry disease (FD) is an X-linked lysosomal disorder with ventricular myocardial involvement that drives morbidity and mortality. Early diagnosis of cardiac involvement can be difficult. This study explored whether abnormal left atrial (LA) strain by cardiovascular magnetic resonance (CMR) may be an early sign of ventricular involvement in FD.

**Methods:** A multicenter, multinational cohort of FD patients was assembled with images centralized for corelab analysis. Adult gene-positive FD patients and healthy volunteers (HV) underwent CMR. LA strain analyses included manually contouring the LA in end-diastole and end-systole to calculate LA volumes and ejection fraction, then semi-automatic analysis for LA reservoir strain.

**Results:** There were n=214 FD patients (mean age 45±15 years, 39% males) and n=76 HV (49±15 years, 53% males). CMR results in FD: LVEF 73% (IQR=9), LV mass indexed (LVMi) 89±39g/m2, 99 (46%) had left ventricular hypertrophy (LVH), 36% had late gadolinium enhancement. In FD, LA strain correlated with LVMi (r=-0.52, p<0.01), LV global longitudinal strain (GLS) (r=-0.61,p<0.01), and native myocardial T1 (r=0.34, p<0.01). FD had abnormal LA strain in overt disease (LVH +ve) compared to HVs (p<0.01). LVH-negative FD did not differ in LA strain compared with HV (p>0.5). FD with low T1+LVH-negative did not differ in LA strain compared with normal T1/LVH-negative FD or HV (p>0.3).

**Conclusions:** LA strain is abnormal in FD with LVH (overt disease) and correlates with LVMi, native T1, and GLS. LA strain is normal in FD with early disease (LVH negative+low T1) and normal in FD with no myocardial disease (LVH negative + normal T1). These findings indicate that LA strain is a consequence of abnormal LV mechanics such as LVH and abnormal GLS, rather than isolated myocardial sphingolipid deposition.

**KEY MESSAGES:** *What is already known on this topic:* Fabry disease is an X-linked lysosomal disorder with potential cardiac complications. Progressive ventricular myocardial involvement drives morbidity and mortality and can be detectable through advanced imaging techniques like cardiovascular magnetic resonance. Early diagnosis of cardiac involvement can be difficult.

*What this study adds:* LA strain is abnormal in overt Fabry disease with left ventricular hypertrophy (LVH) and correlates with left ventricular mass, native T1, and global longitudinal strain (GLS). LA strain is normal in Fabry with early disease (LVH negative+low T1) and normal in FD with no myocardial disease (LVH negative + normal T1). These findings indicate that LA strain is a consequence of abnormal LV mechanics such as LVH and abnormal GLS, rather than isolated myocardial sphingolipid deposition.

*How this study might affect research:* There is a need to identify markers of early cardiac involvement in Fabry disease.

## INTRODUCTION

Fabry disease (FD) is a rare condition caused by a mutation in the GLA gene resulting in deficiency of the enzyme alpha-galactosidase A.^1^ Although FD is X-linked, it affects both men and women.^2^ Cardiac involvement drives mortality and morbidity,^5,6^ and is thought to be due to sphingolipid deposition in cardiac myocytes and vascular endothelium, with associated inflammation, hypertrophy, and fibrosis leading with time to increased arrhythmias, heart failure and sudden death. ^3,4^ However, all components of the heart may be affected, including the atria and valves. ^26^

Left Atrial (LA) function, known as left atrial strain – via its reservoir, conduit, and contractile phases during the cardiac cycle – is heavily influenced by atrial compliance + contractility. ^18,19^ Abnormal LA strain in FD has been shown to be associated with cardiac complications including atrial fibrillation and stroke, as well as ventricular structural changes such as left ventricular hypertrophy (LVH). ^4,7,21,22^ Abnormal atrial strain has been documented with echocardiography prior to the development of LVH.^22^

Cardiovascular magnetic resonance (CMR) adds value to the detection of key cardiovascular processes in the FD phenotype. CMR can detect LVH/cardiac mass with increased precision, and key is its ability to characterise myocardial tissue with late gadolinium enhancement (LGE, scar/inflammation) and more recently T1 mapping. Low T1 is a surrogate marker of myocardial sphingolipid deposition, which is a marker of early disease.^13^ ^3,4^ CMR has also been useful in proposing the different stages of Fabry disease, for example the pre-hypertrophy stage^8,28^.

Our hypothesis was that abnormal LA strain is an early sign of cardiac involvement in Fabry disease. Thus, the aim of this study was to investigate left atrial strain by CMR in Fabry disease and its association with low myocardial native T1, LVH and other CMR markers of Fabry disease.

## METHODS

This study is a secondary analysis of a prospective, multicentre international observational study (Fabry400, NCT03199001), which recruited patients with Fabry disease from 3 sites (London, Birmingham, and Sydney).

The study was approved by the relevant local Research Ethics Committees (eg, Northern Sydney Local health District Human Research Ethics Committee), and written informed consent was obtained from all participants. The inclusion criteria were any patient with gene-positive Fabry disease and adults ≥18 years. Healthy volunteers were all prospectively recruited volunteers from a single site with no history of cardiovascular disease (normal health questionnaire, normal electrocardiogram, no cardioactive medication unless for primary prevention).

### CMR imaging

All participants underwent CMR at 1.5 Tesla (Avanto (UK), Aera (Australia); Siemens Healthcare, Erlangen, Germany) using a standard protocol, including cines, T1 mapping (MOLLI), and late gadolinium enhancement imaging, as previously published. ^27^

### CMR analysis

Conventional CMR parameters had already been analysed and published on both FD and HV cohorts (eg LV volumes, function, global longitudinal strain (GLS), mass, LGE (presence/absence) and native myocardial T1)^14^. For this study, a new analysis was performed of Left atrial (LA) strain. This used CVI42 software (Circle Cardiovascular Imaging Inc., Calgary, Canada) and included manually contouring the LA in end diastole and end systole in both the 4-chamber and 2-chamber views to derive LA maximum and minimum volumes. These were then interpolated throughout the cardiac cycle by the software (with manual correction when needed) to derive the analysis of atrial function including LA ejection fraction, and LA strain (reservoir strain). To improve precision, the analyses were repeated five times for each patient and the derived LA strain result was the mean of the 5 analyses, ‘mean reservoir strain’.

Intra-observer and inter-observer repeatability of left atrial strain were also analysed and reported (n=10) random subset).

### Statistical Analysis

Statistical analyses were carried out using SPSS (IBM, Armonk, NY). Continuous variables are expressed as mean ± standard deviation, categorical as frequencies or percentages. Normality was checked using the Shapiro-Wilk test. Groups were compared using the independent-samples t-test (normally distributed variables) or the Mann-Whitney U test (non-normally distributed). ANOVA was used to assess significant differences between groups and regression models were used to assess the relationships between LA strain, T1 values and LVH status. A p-value of <0.05 was considered statistically significant. A Bland-Altman analysis was carried out to calculate the intra and interobserver variability and 95% confidence intervals.

## RESULTS

### Participant Characteristics

There were 290 participants in total (214 FD and 76 HV), all in sinus rhythm. Of note, eight patients were excluded from the LA strain statistical analysis prior to the total population count, due to either incomplete phase scans, or poor image quality. Adequate tracking quality was obtained for all other study participants (n=290). Baseline participant demographics and CMR findings are demonstrated in Table 1. The mean Fabry age was 45±15 years with 83(39%) males and 131(61%) females. The HV population were slightly older, mean age (49±14 years, p=0.01; females 48% and males 52%).

**Table 1.**
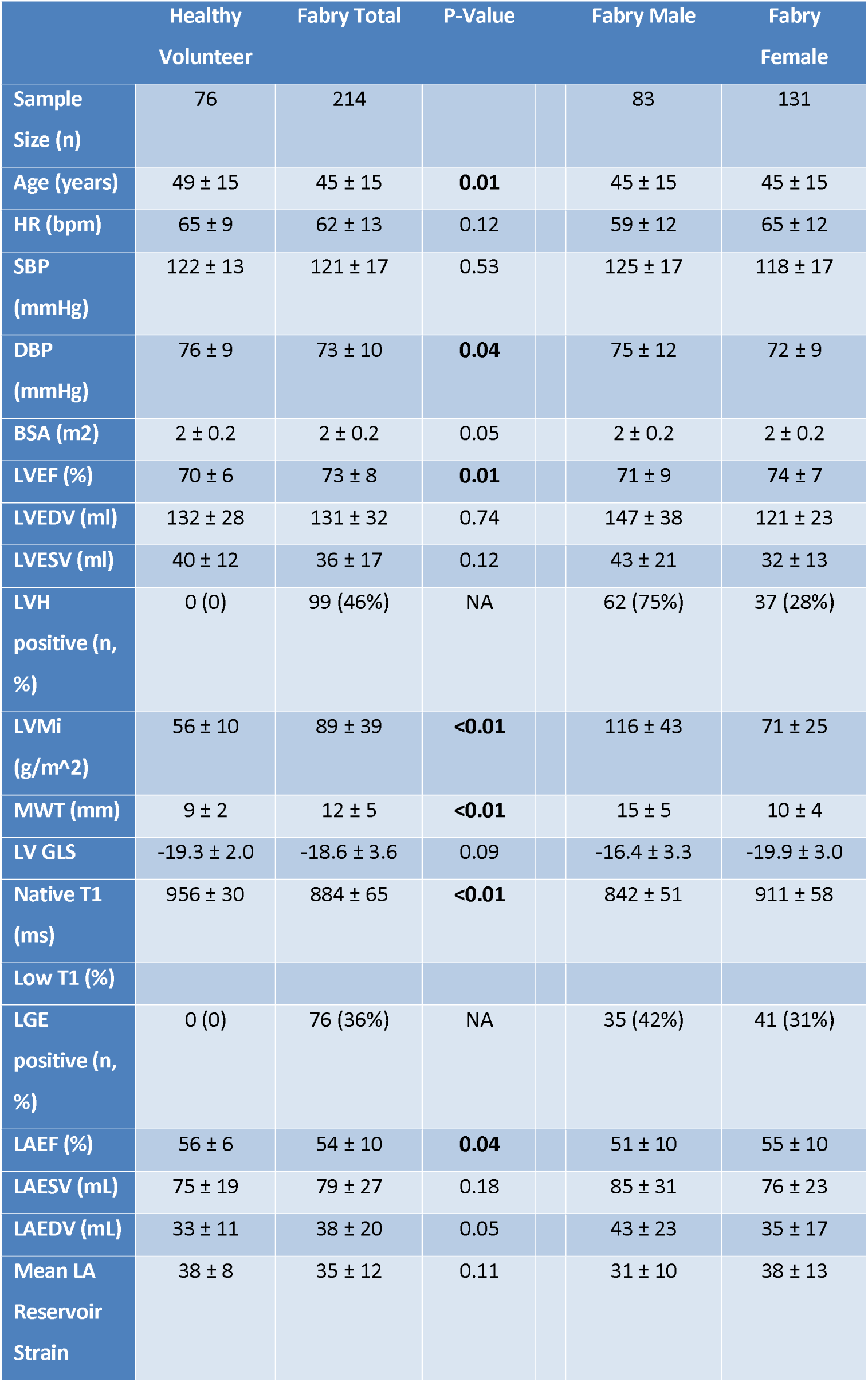
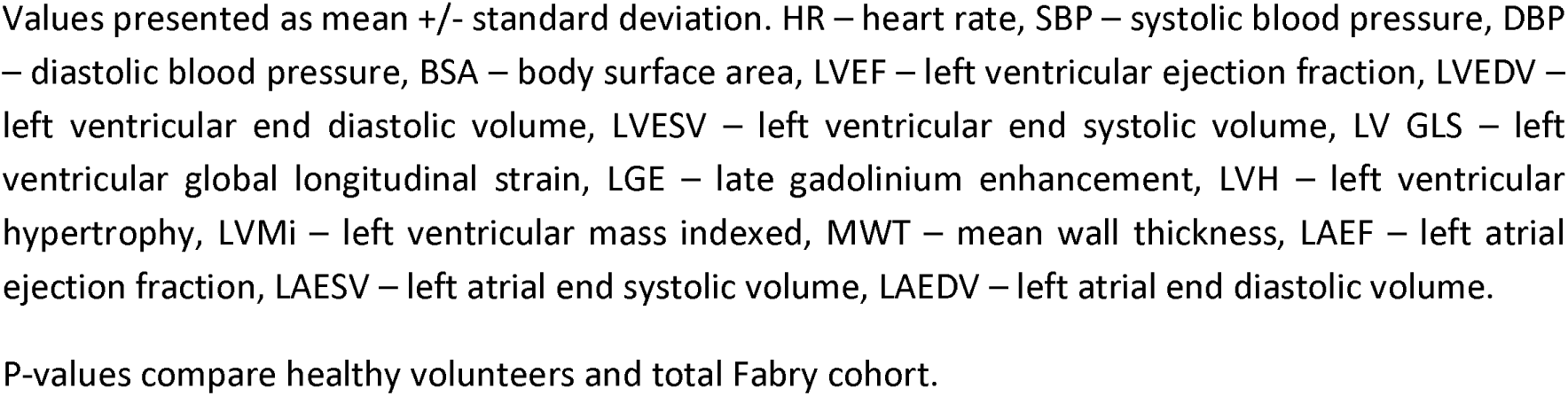
Participant demographics and basic CMR findings. Values presented as mean +/- standard deviation. HR – heart rate, SBP – systolic blood pressure, DBP – diastolic blood pressure, BSA – body surface area, LVEF – left ventricular ejection fraction, LVEDV – left ventricular end diastolic volume, LVESV – left ventricular end systolic volume, LV GLS – left ventricular global longitudinal strain, LGE – late gadolinium enhancement, LVH – left ventricular hypertrophy, LVMi – left ventricular mass indexed, MWT – mean wall thickness, LAEF – left atrial ejection fraction, LAESV – left atrial end systolic volume, LAEDV – left atrial end diastolic volume. P-values compare healthy volunteers and total Fabry cohort.

|  | Healthy Volunteer | Fabry Total | P-Value |  | Fabry Male | Fabry Female |
| --- | --- | --- | --- | --- | --- | --- |
| Sample Size (n) | 76 | 214 |  |  | 83 | 131 |
| Age (years) | 49 ± 15 | 45 ± 15 | **0.01** |  | 45 ± 15 | 45 ± 15 |
| HR (bpm) | 65 ± 9 | 62 ± 13 | 0.12 |  | 59 ± 12 | 65 ± 12 |
| SBP (mmHg) | 122 ± 13 | 121 ± 17 | 0.53 |  | 125 ± 17 | 118 ± 17 |
| DBP (mmHg) | 76 ± 9 | 73 ± 10 | **0.04** |  | 75 ± 12 | 72 ± 9 |
| BSA (m2) | 2 ± 0.2 | 2 ± 0.2 | 0.05 |  | 2 ± 0.2 | 2 ± 0.2 |
| LVEF (%) | 70 ± 6 | 73 ± 8 | **0.01** |  | 71 ± 9 | 74 ± 7 |
| LVEDV (ml) | 132 ± 28 | 131 ± 32 | 0.74 |  | 147 ± 38 | 121 ± 23 |
| LVESV (ml) | 40 ± 12 | 36 ± 17 | 0.12 |  | 43 ± 21 | 32 ± 13 |
| LVH positive (n, %) | 0 (0) | 99 (46%) | NA |  | 62 (75%) | 37 (28%) |
| LVMi (g/m<sup>2</sup>) | 56 ± 10 | 89 ± 39 | **<0.01** |  | 116 ± 43 | 71 ± 25 |
| MWT (mm) | 9 ± 2 | 12 ± 5 | **<0.01** |  | 15 ± 5 | 10 ± 4 |
| LV GLS | -19.3 ± 2.0 | -18.6 ± 3.6 | 0.09 |  | -16.4 ± 3.3 | -19.9 ± 3.0 |
| Native T1 (ms) | 956 ± 30 | 884 ± 65 | **<0.01** |  | 842 ± 51 | 911 ± 58 |
| Low T1 (%) |  |  |  |  |  |  |
| LGE positive (n, %) | 0 (0) | 76 (36%) | NA |  | 35 (42%) | 41 (31%) |
| LAEF (%) | 56 ± 6 | 54 ± 10 | **0.04** |  | 51 ± 10 | 55 ± 10 |
| LAESV (mL) | 75 ± 19 | 79 ± 27 | 0.18 |  | 85 ± 31 | 76 ± 23 |
| LAEDV (mL) | 33 ± 11 | 38 ± 20 | 0.05 |  | 43 ± 23 | 35 ± 17 |
| Mean LA Reservoir Strain | 38 ± 8 | 35 ± 12 | 0.11 |  | 31 ± 10 | 38 ± 13 |

Compared to HV, CMR results showed that FD had higher LVEF (73±8% vs 70±6%, p=0.01) but similar LV GLS (p=0.09). Mean indexed LV mass (LVMi) was higher in FD compared to HV (89±39g/m^2^ vs 56±10g/m^2^, p<0.001). 99(46%) of FD had LVH (males 62/83 (75%), females 37/131 (28%)). No HV had LVH. 72% of FD had low T1 and no HV had low T1. 36% of FD had LGE.

### Left Atrial Function

Compared to HV, FD had similar LA end systolic volumes (FD 79.2 ± 26.9 ml vs HV 74.6 ± 18.9 ml, p=0.18), but larger LA end diastolic volumes (FD 37.6 ± 20.0 ml, vs HV 32.9 ± 10.9 ml, p=0.05) and lower LA ejection fraction (FD 53.8 ± 10.2, vs HV 56.4 ± 6.2, p=0.04).

Table 2 displays the LA strain values in HV and FD patients. Overall, LA reservoir strain was similar in FD compared to HV (35.2 ± 12.0 % versus 37.6 ± 8.3 %, p=0.06). However, when compared according to sex, male FD had worse LA reservoir strain compared to male HV (34.8 ± 7.4 % versus 30.7 ± 9.7 %, p=0.01) and female FD had similar LA reservoir strain compared to female HV (40.6 ± 8.3 % versus 38.0 ± 12.5 %, p=0.145).

**Table 2.** mean LA reservoir strain in subgroups. *P values are comparing Fabry to healthy volunteers.

|  |  | HV LA Strain |  | Fabry LA Strain |  | P values |
| --- | --- | --- | --- | --- | --- | --- |
| | | N | Mean ± SD | N | Mean ± SD | |
| Study population | Total | 76 | 37.6 ± 8.3 | 214 | 35.2 ± 12.0 | 0.069 |
| | Male | 40 | 34.8 ± 7.4 | 83 | 30.7 ± 9.7 | **0.010** |
| | Female | 36 | 40.6 ± 8.3 | 131 | 38.0 ± 12.5 | 0.145 |
| LVH-positive | Total | | | 99 | 29.2 ± 9.6 | **<0.001** |
| | Male | | | 62 | 29.5 ± 9.8 | **0.003** |
| | Female | | | 37 | 28.7 ± 9.4 | **<0.001** |
| LVH-negative | Total | | | 115 | 40.3 ± 11.5 | 0.052 |
| | Male | | | 21 | 34.3 ± 8.6 | 0.822 |
| | Female | | | 94 | 41.7 ± 11.6 | 0.549 |

\*P values are comparing Fabry to healthy volunteers.

Figure 1 shows the relationship between LA strain (reservoir strain) in the total FD cohort and structure (LVM), function (LVEF, LV GLS) and tissue characterisation (native T1).

**Figure 1.**
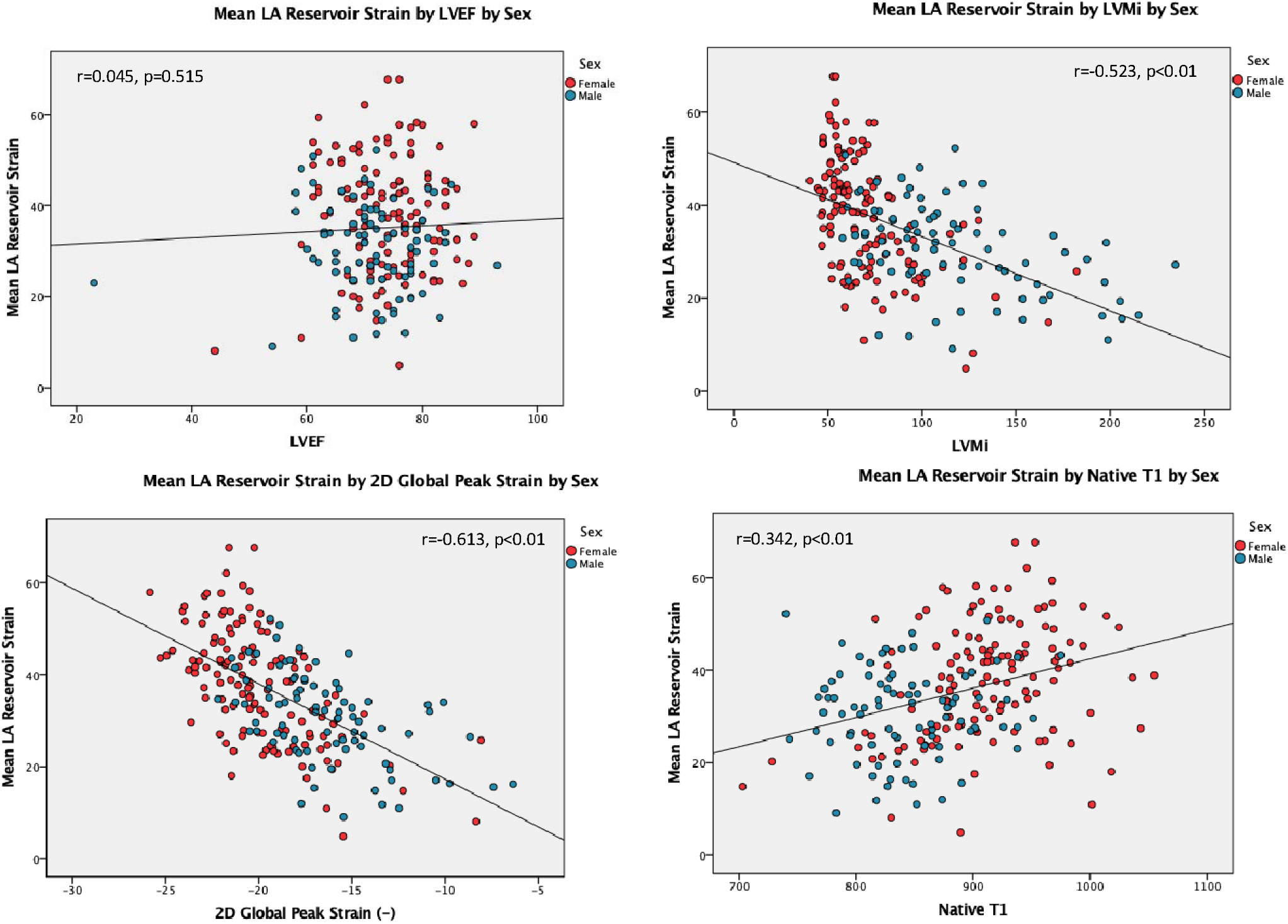
plots showing relationship between mean LA reservoir strain and left ventricular ejection fraction (LVEF), left ventricular mass index (LVMi), left ventricular global longitudinal strain (GLS) and T1.

### Left atrial strain & left ventricular mass

LA strain correlated with LVMi in the total Fabry cohort (as LVMi increased LA strain worsened, (r=-0.52, p<0.01). Table 2 shows the LA reservoir strain measurements in HV and FD divided into males and females and also those with and without LVH. FD with LVH (overt disease) had worse LA strain compared to FD without LVH and HV (29.2 ± 9.6 vs 40.3 ± 11.5 vs 37.6 ± 8.3, p<0.001) and this was also seen in both males and females. FD without LVH had similar LA strain compared with HV both in males (34 ± 8.6 vs 34.8 ± 7.1, p=0.82) and females (41.7 ± 11.6 vs 40.6 ± 8.3, p=0.55).

### Left atrial strain & left ventricular function

LA strain did not correlate with LVEF but it did correlate with LV GLS (as GLS worsened so did LA strain, (r=-0.61, p<0.01). Table 3 shows LA strain in patients with abnormal GLS with and without LVH. Normal GLS was determined as the mean ± 2 standard deviations in healthy volunteers (19.3 ± 2.0%). In patients with abnormal GLS, LA strain was significantly impaired in LVH compared to no LVH (27.1 ± 9.5% vs 41.6 ± 12.3%, p<0.001, normal LA strain 37.6 ± 8.3%). LA strain was also impaired in patients with LVH but normal GLS (32.6±8.2%).

**Table 3.** Differences in mean LA reservoir strain values between GLS and LVH status.

|  | N | LVH negative | N | LVH positive | p-value |
| --- | --- | --- | --- | --- | --- |
| Normal GLS | 63 | 40.0±12.0 | 37 | 32.6±8.2 | **p<0.001** |
| Abnormal GLS | 53 | 41.6±12.3 | 63 | 27.1±9.5 | **p<0.001** |
|  |  | **p<0.001** |  | **p<0.001** |  |

### Left atrial strain & native T1

LA strain correlated with native T1 (as native T1 lowered LA strain worsened, (r=0.34, p<0.01). Table 4 shows LA strain in FD with no LVH + low T1 (early disease) and FD with no LVH + normal T1 (no myocardial sphingolipid deposition). FD with no LVH + low T1 had similar LA strain to HV. FD with no LVH + normal T1 had supra-normal LA strain compared to HV, but when divided by sex, there were no significant differences between these subgroups. Interobserver coefficient of repeatability = 10.20

**Table 4.** mean LA reservoir strain values in LVH-negative Fabry population classified according to native T1 and compared with healthy volunteers.

|  | HV |  | LVH-negative Fabry + low T1 |  |  | LVH-negative Fabry + normal T1 |  |  |
| --- | --- | --- | --- | --- | --- | --- | --- | --- |
|  | N | Mean±SD | N | Mean±SD | P value | N | Mean±SD | P value |
| **Total** | 76 | 37.6 ± 8.3 | 64 | 39.0 ± 10.3 | 0.351 | 51 | 42.0 ± 12.7 | **0.029** |
| **Male** | 40 | 34.8 ± 7.4 | 16 | 34.9 ± 9.2 | 0.969 | 5 | 32.4 ± 6.5 | 0.476 |
| **Female** | 36 | 40.6 ± 8.3 | 48 | 40.4 ± 10.3 | 0.922 | 46 | 43.0 ± 12.8 | 0.308 |

\*P values are comparing Fabry to healthy volunteers

### Reproducibility, Intra- and inter-observer variability

Assessment of the intra- and inter-observer variability showed acceptable levels of agreement for LA reservoir strain. Figure 2 shows the Bland Altman graphs and coefficients of repeatability.

**Figure 2.**
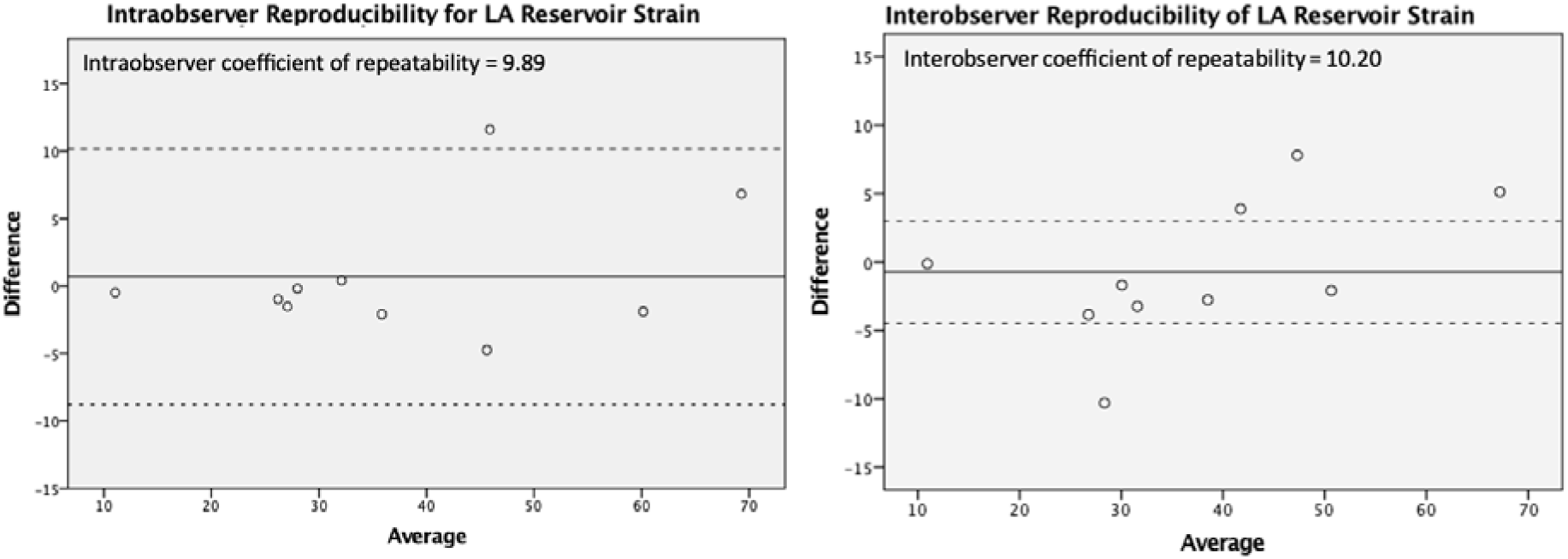
Bland Altman graphs for LA strain intraobserver (left) and interobserver (right) repeatability.

## DISCUSSION

The main finding of this CMR study is that LA strain is abnormal in FD patients with LVH but normal in FD with no LVH. Additionally, LA strain correlates with increased LVMi, low native T1, and abnormal GLS in FD. In LVH- negative Fabry patients, i.e. those with ‘early disease’, LA strain does not correlate with native T1. These findings indicate that LA strain is not influenced solely by sphingolipid deposition, but rather as a consequence of structural changes such as LVH and LV GLS. Therefore, abnormal LA strain by CMR can be used as an imaging marker of cardiac involvement.

Regarding how these results compare to the existing literature, there is a notable lack of consensus and ongoing debate in the literature regarding whether LA dysfunction in Fabry disease is caused by the progression of ventricular dysfunction, or rather, is caused by sphingolipid deposition in the LA itself.^7^ One theory states that the build-up of sphingolipids in the LA myocardium results in interstitial fibrosis, which affects LA size and causes mechanical dysfunction, ultimately putting the patient at risk of AF.^7^ The alternate theory suggests that deposition of sphingolipids in the LV may cause decreased LV compliance, diastolic dysfunction and higher LV filling pressures. The thin LA wall is particularly sensitive to changes in LV filling pressures, and so this dysfunction is then passively transmitted to the LA.^7^

A recent study showed that in LVH-negative FD, impairment of left ventricular global longitudinal strain by CMR was associated with a reduction in native T1.^14^ These findings suggests that left ventricular mechanical dysfunction may occur before evidence of LVH, when there is evidence of low T1 (sphingolipid deposition).^14^ However, to date most studies of LA strain in Fabry patients have been conducted on small populations using echocardiography speckle-tracking imaging, rather than CMR.^24,25^ Phasic LA function, particularly conduit and reservoir LA strain, measured using MRI is independently predictive of heart failure hospitalisations and death in other (non-Fabry) patient populations.^25^ Thus there is an important gap in the current literature regarding CMR investigation of LA function in Fabry disease, which this study addresses.

Recent studies by Boyd et al. have argued that LA dysfunction occurs before the development of LVH in Fabry patients.^22^ Their study used echocardiography speckle-tracking to show that increased LA volume and reduced atrial compliance occurred in Fabry patients in the absence of LVH. Boyd et al. therefore argued that Fabry disease alters atrial myocardial properties early in the disease process, and that LA dysfunction is therefore largely due to atrial myocardial fibrosis. Similarly, further echocardiography investigation from Pichette et al. suggests that sphingolipids also accumulate in the LA, and that LA dysfunction is therefore not just due to passive transmission of LV dysfunction.^21^ Conversely, Saccheri et al. found that LA dysfunction and changes to atrial volume in Fabry disease are likely associated with the progression of LV dysfunction, rather than the deposition of sphingolipids in the LA myocardium.^7^ The results of our study support the latter, that LA dysfunction is largely a consequence of LV changes such as LVH and abnormal GLS, and sits on the continuum of ventricular storage to left ventricular hypertrophy.

There are several limitations identified in this project. Firstly, Fabry disease is very rare, and as such any studies on the subject are inherently limited in their sample sizes, and therefore the statistical power of any results obtained. Considering this, this study represents one of the largest Fabry datasets to date (n=214). However, it is a retrospective study that included patients already diagnosed with Fabry disease, potentially introducing bias, and there are small numbers when comparing subgroups, particularly males with no LVH. Secondly, there are difficulties in standardising the analyses of LA strain using CMR, as there are differences depending on the software used (CVI42 in this study), and differences when compared to echocardiography analyses. There are other challenges associated with strain imaging of the left atrium that may undermine its diagnostic accuracy^17^ such as the thin LA wall and the confounding presence of the atrial appendage and pulmonary veins, which complicate the measurement of LA strain parameters using the current technology.^18,19^ Thirdly, this study did not look at confounding effects of co-morbidities, such as renal disease, nor disease specific therapy (e.g. enzyme replacement therapy). Fourthly, it is difficult to comment on sex differences of LA strain in patients with Fabry disease. In our cohort, females had smaller LVEDV, lower LVMi and normal mean wall thickness ie, less left ventricular hypertrophy (75% in males and 28% in females). The results of our study suggest that LA strain becomes abnormal when LVH occurs, hence it was not surprising that females had normal mean LA strain compared to males and healthy controls because they had less LVH. In order to better explore sex differences in LA strain in Fabry disease, future studies need to include more females with LVH. Finally, this study is limited by the absence of histology confirmation of the extent of sphingolipid deposition and fibrosis within the LV and LA. Consequently, asserting that the subgroup displaying no LVH and normal T1 values represents individuals with no cardiac involvement of Fabry disease, and patients with no LVH and low T1 values represent very early disease, although plausible, cannot be definitively stated.

## CONCLUSIONS

LA strain is abnormal in FD with LVH (overt disease) and correlates with LVMi, native T1, and GLS. LA strain is normal in FD with early disease (LVH negative + low T1) and normal in FD with no myocardial disease (LVH negative + normal T1). These findings indicate that LA strain is a consequence of abnormal LV mechanics such as LVH and abnormal LV GLS rather than isolated myocardial sphingolipid deposition.

## Data Availability

All data produced in the present work are contained in the manuscript.

## ACKNOWLEDGMENTS

This study is part of the Fabry400 study (NCT03199001), which was funded by an investigator led research grant from Sanofi-Genzyme.

## DATA AVAILIBILITY STATEMENT

The data underlying this article will be shared on reasonable request to the corresponding author.

## ABBRIEVIATIONS LIST

CMR: cardiovascular magnetic resonance
FD: Fabry disease
GLS: global longitudinal strain
HV: healthy volunteers
LA: left atrium
LGE: late gadolinium enhancement
LVMi: left ventricular mass index
LVH: left ventricular hypertrophy
LV: left ventricle
LVEF: left ventricular ejection fraction

